# The phenomic landscape of rare gain- and loss-of-function variation

**DOI:** 10.64898/2026.11.11.26367788

**Authors:** Meltem Ece Kars, David Stein, Yiming Wu, Zsu-Zsu Chen, Shuliang Deng, Hanna Abe, Peter D. Stenson, David N. Cooper, Laura M. Raffield, Leslie A. Lange, Michael Tsai, Mark O. Goodarzi, Stephen S. Rich, Jerome I. Rotter, Paul Cohen, Robert E. Gerszten, Yuval Itan

## Abstract

Current large-scale phenome-wide association studies do not distinguish between genetic variants with distinct functional consequences, despite evidence that gain-of-function (GoF) and loss-of-function (LoF) variants can produce different phenotypes. Consequently, the phenotypic consequences of variants stratified by predicted GoF and LoF effects have not been systematically characterized at scale. Here, we systematically analyzed rare predicted GoF and LoF variants in 18,214 genes across 1,244 binary and 202 quantitative traits using whole-exome sequencing and electronic health records from the Mount Sinai Million Health Discoveries Program, a phenotypically rich hospital-based biobank comprised of 50,067 participants, primarily representing African, Admixed American and European ancestries. We identified numerous established and previously unreported ancestry-specific and shared gene– and variant-level associations, many of which were supported by complementary evidence from both diagnostic phenotypes and clinical measurements. Proteomic evaluation of the previously unreported association between *VNN1* LoF variants and body mass index in the Multi-Ethnic Study of Atherosclerosis revealed reduced circulating vanin-1 levels with LoF variants, which in turn were associated with body composition and additional cardiometabolic phenotypes. Functional stratification of predicted GoF and LoF variants revealed gene-trait associations that were attenuated when these variants were combined, demonstrating that traditional aggregation of variants with different functional effects can obscure association signals. These findings underscore the importance of incorporating predicted functional consequences into association analyses and establish a comprehensive resource linking exome-wide predicted GoF and LoF variants to the human phenome across ancestry-specific and multi-ancestry analyses.

## Main

Investigating the functional effect of genetic variants on human phenotypes has been the central focus of human disease genomics. Variants in the coding region of the genome can influence molecular binding, stability, catalytic activity, and other biochemical properties of proteins, thereby leading to various functional consequences. The two major categories of functional consequences are gain-of-function (GoF), which is characterized by a new or enhanced protein function, and loss-of-function (LoF), which causes either a partial or complete loss of protein activity.

Traditionally, investigation of novel disease genes and variants is performed using general variant pathogenicity predictors without taking the GoF and LoF effects into account^1^. However, because of their distinct impact on proteins, GoF and LoF variants in the same gene can result in different phenotypes. For example, LoF variants in *RET* cause Hirschsprung’s disease^2^, characterized by loss of motility in the distal colon due to lack of neural crest cells, whereas GoF variants in *RET* lead to multiple endocrine neoplasia type 2^3^, a tumor syndrome of neural crest origin affecting multiple tissues. Therefore, gaining further insight into diverse phenotypic consequences of GoF and LoF variants could significantly enhance our understanding of disease pathophysiology and development of treatment strategies. We recently developed LoGoFunc, a machine learning variant classifier of GoF and LoF missense variants, trained on a broad range of gene-, protein-, and variant-level features describing diverse biological characteristics^4^. LoGoFunc outperforms other tools trained solely on predicting pathogenicity by distinguishing GoF and LoF effects. Using LoGoFunc, we obtained GoF, LoF and neutral prediction scores for all possible human missense variants and demonstrated that utilizing these scores facilitates the identification of disease associated variants and genes, as well as candidate therapeutic targets.

Phenome-wide association studies (PheWAS) have been instrumental in investigating genotype-phenotype correlations in large scale biobanks. Adopting a genotype-first approach, PheWAS enables hypothesis-free exploration of disease associations in deeply-phenotyped biobanks that include electronic health records (EHR) and genetic data. Recently, several large-scale PheWAS resources have been publicly released, including those that utilized UK Biobank^5,6^, Veterans Affairs Million Veteran Program (MVP)^7^ and All of Us Research Program^8,9^ datasets. These resources differ in their methodologies (gene– and/or variant-based association testing), genetic data sources (rare variants identified using exome or whole genome sequencing versus genotyping array of common variants), phenotype definitions (ICD– or phecode-based binary traits versus quantitative traits obtained from laboratory measurements) and ancestral composition (European-predominant or multiple population groups). However, there is still an unmet need for large-scale analyses of rare variants to identify disease-associated genetic factors in phenotypically well-characterized and diverse population cohorts. Moreover, the differential effects of GoF and LoF variants across a broad range of phenotypes have yet to be investigated.

Here, we describe a systematic, large-scale gene– and single variant-level analysis of rare GoF and LoF variants in 50,067 participants from the Mount Sinai Million Health Discoveries Program (MSM), a deeply phenotyped and genetically diverse hospital-based biobank that links whole exome sequencing (WES) data with EHR from the Mount Sinai Health System in New York City (**Fig. 1**). Furthermore, we performed a focused proteomic evaluation of the previously unreported association between *VNN1* LoF variants and body mass index in the Multi-Ethnic Study of Atherosclerosis (MESA). Our findings underscore the significance of distinguishing the functional impact of genetic variants in association studies and emphasize the importance of expanding PheWAS to populations from diverse genetic ancestries and to quantitative traits, which can both support findings from analyses of binary traits and facilitate the identification of novel clinical associations. To accelerate these efforts, we generated a publicly accessible resource of exome-wide PheWAS results for predicted GoF and LoF variants and genes across 1,446 phenotypes, including ancestry-specific and multi-ancestry analyses.

**Fig. 1:**
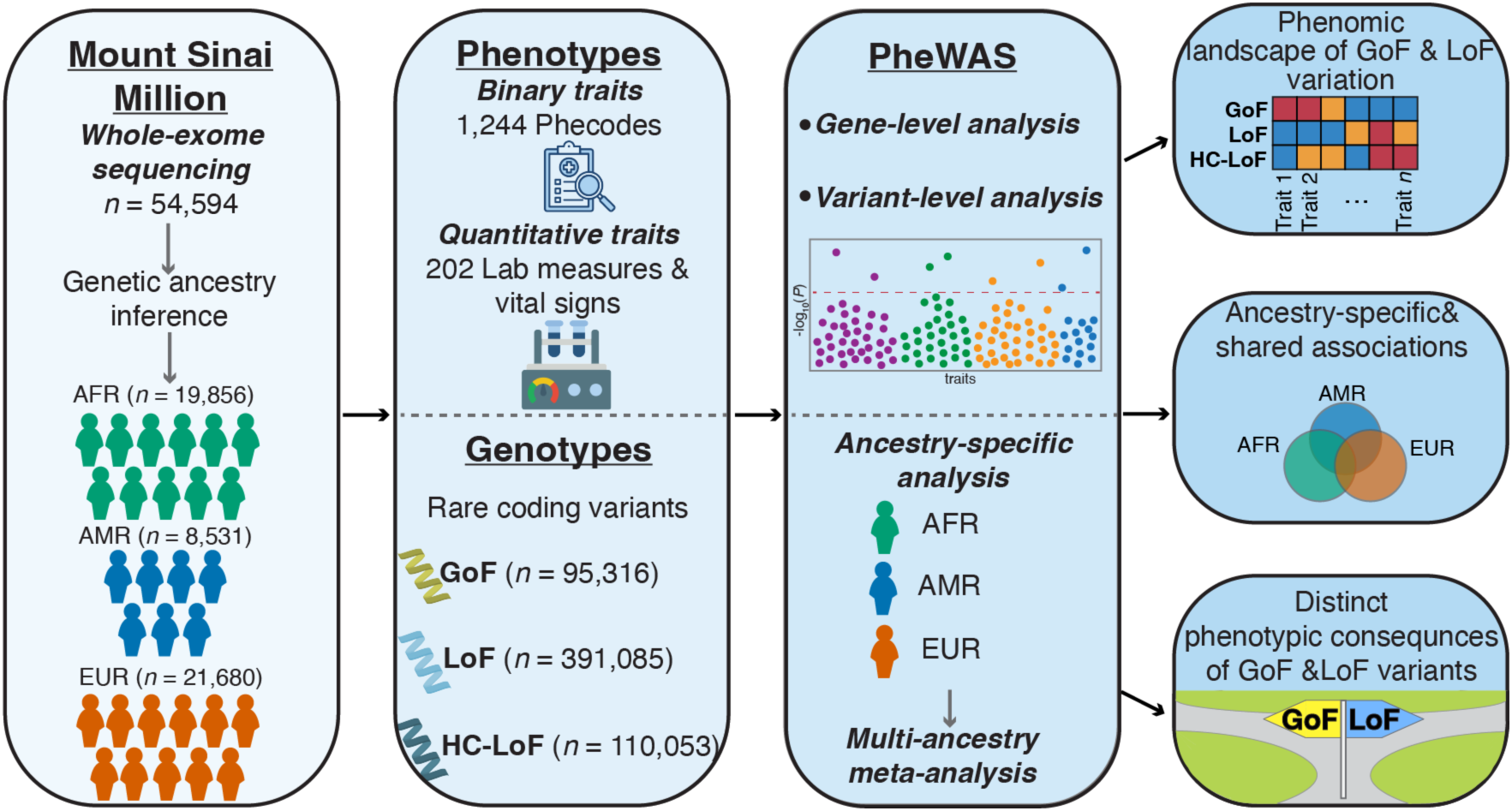
Study workflow. Schematic overview of the study design, illustrating the study cohorts, phenotype curation and variant selection, association testing methods, and major conclusions from left to right.

## Study cohort and phenotypes

MSM contains whole exome sequencing (WES) data linked to EHR of study participants admitted to the Mount Sinai health system. The genetically inferred ancestries of 58,990 high-quality samples were determined based on genetic similarity to HapMap3 populations^10^. This included African (AFR, *n* = 19,856), Admixed American (AMR, *n* = 8,531), East Asian (EAS, n = 2,464), European (EUR, *n* = 21,680), and South Asian (SAS, n = 2,063) populations (**Fig. 2a and Extended Data Fig. 1**). Due to the relatively small sample sizes of the EAS and SAS groups, we included 50,067 MSM samples from the AFR, AMR, and EUR ancestry groups in the study. The cohorts predominantly consisted of adult participants, with a mean age of 61.6 in AFR and EUR, and 58.8 in AMR. The proportion of females was slightly higher than that of males across the population groups (**Fig. 2b & Supplementary Table 1**).

**Fig. 2:**
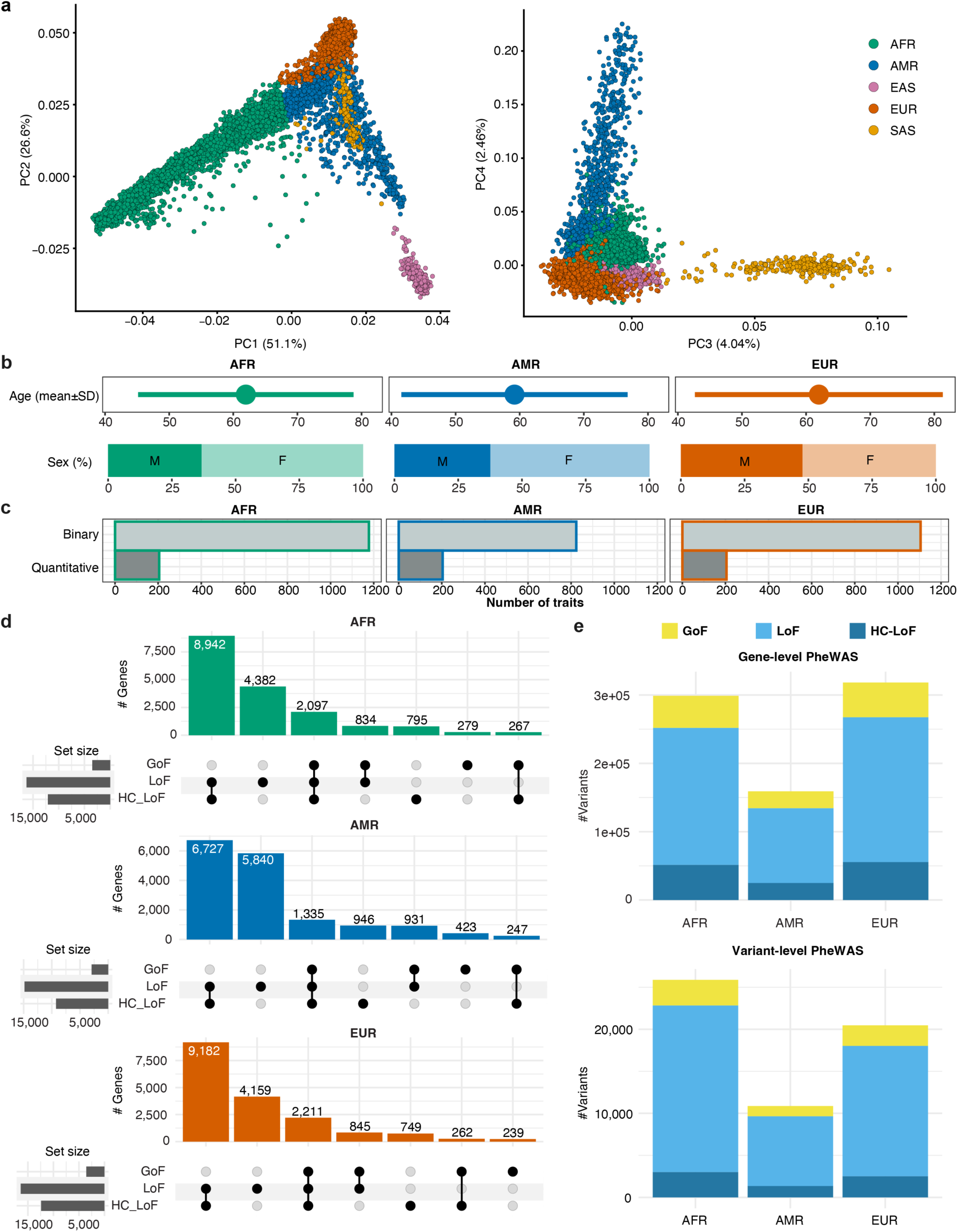
Demographic characteristics of the study cohorts and variant summary. **a**, Principal components 1 and 2 (left) and 3 and 4 (right) demonstrating the distribution of genetic ancestries of MSM participants (*n* = 58,990). **b,** Distribution of age and sex across ancestry-specific cohorts (AFR, *n* = 19,856; AMR, *n* = 8,531; and EUR, *n* = 21,680). **c,** Number of binary and quantitative phenotypes in each ancestry-specific cohort. **d,** UpSet plot depicting the number of genes harboring different variant categories. **e,** Number of GoF, LoF and HC-LoF variants included in the gene– and variant-level analyses.

We processed ICD-9 and ICD-10 diagnoses of MSM participants by mapping them to 3,200 phecodes based on phecodeX definitions^11^ and included phecodes with at least 50 cases in each population group corresponding to a total of 1,244 binary traits. Additionally, we curated a set of 202 laboratory measurements and vital signs from EHR data and calculated median, minimum, and maximum values for each participant. These quantitative traits were observed in at least 50 participants in each population group (**Fig. 2c, Supplementary Tables 1 and 2**).

We compared the prevalence of binary traits across MSM population groups and found that, although the EUR cohort had the largest sample size, the AFR cohort had 137 traits not included in the EUR analysis due to small sample sizes (*n* < 50), and 55% of 1,041 traits tested in both AFR and EUR were at least 1.5-fold more prevalent in AFR. Similarly, comparisons of the AMR and EUR cohorts showed that AMR had 14 traits not included in the EUR analysis, and 47% of 810 traits tested in both cohorts were at least 1.5-fold more prevalent in the AMR cohort (**Supplementary Table 2**). Additionally, the AFR and AMR cohorts exhibited higher prevalence across most phecode categories compared with the EUR cohort (**Extended Data Fig. 2**). Although disease prevalence in a hospital-based biobank does not directly reflect its prevalence in the general population and is influenced by disparities in health and healthcare access, these findings underscore the importance of expanding genetic association studies to non-EUR populations, as well as conducting population-specific association studies.

## Identifying rare predicted GoF and LoF variants

After processing WES data from 50,067 participants and performing quality control (QC) at the variant and genotype levels, we identified 4,900,775 high-quality variants in 24,174 genes. We then extracted variants with a minor allele frequency (MAF) <1% in all gnomAD v4.0.1 population groups^12^ and identified 95,316 GoF and 391,085 LoF rare missense variants based on LoGoFunc predictions. Additionally, we identified 110,053 rare protein truncating variants, defined as high-confidence LoF (HC-LoF) variants by LOFTEE^13^, including frameshift, stop-gained and essential splice site variants, predicted to have a high impact on gene products. Overall, we identified a total of 596,454 rare GoF, LoF and HC-LoF variants in 18,214 genes.

The number of genes harboring at least one LoF or HC-LoF variant was consistently higher than the number of genes carrying at least one GoF variant across the three ancestry groups. Most genes harbored both LoF and HC-LoF variants, while only 79 genes carried only GoF and 3,680 genes carried both GoF and LoF/HC-LoF variants across the three ancestry groups (**Fig. 2d**). The number of variants observed in each ancestry group was largely in line with the corresponding sample size (**Fig. 2e**). We also extracted 1,808,714 synonymous variants from genes harboring GoF, LoF and HC-LOF variants using the same gnomAD MAF threshold to calculate the genomic inflation factor for each trait under a negative control model. The resulting variant sets were used in downstream gene– and variant-level PheWAS analyses in each ancestry group.

## Establishing high-quality gene– and variant-level PheWAS results

We conducted gene-level PheWAS in each ancestry group using the Scalable and Accurate Implementation of Generalized mixture model (SAIGE) framework^14^, based on 5 variant sets (GoF, LoF, HC-LoF, LoF+HC-LoF and GoF+LoF+HC-LoF) across 1,244 binary and 202 quantitative traits (tested separately using minimum, median and maximum values per sample), corresponding to a total of 284,775,878 set-based association tests. The SAIGE framework also includes a single variant testing option, which we used to perform variant-level PheWAS, corresponding to 72,140,184 variant-level association tests (**Supplementary Table 3**). Given that the analyses encompassed multiple ancestry groups, variant sets and traits with varying case counts, we considered associations with *P* < 1 x 10^-5^ as suggestive associations for exploratory comparisons across ancestry groups and variant sets. We further applied Benjamini-Hochberg false discovery rate (BH-FDR) correction for each phenotype and variant set to identify statistically significant associations (see Methods).

We then conducted both gene– and variant-level PheWAS using rare synonymous variants from the same genes as an internal negative control to perform QC on summary statistics. Based on the distribution of genomic inflation factors (λ) across all tested traits and ancestry groups in the synonymous variant analyses, we observed improved calibration after restricting the gene-level tests to those with a cumulative minor allele count (MAC) ≥ 10 and variant-level tests to those with a MAC ≥ 10. Traits that still exhibited severe deflation or inflation were excluded from the summary statistics (**Extended Data Fig. 3 and 4**). Based on these criteria, we obtained 117,304,896 gene-level and 42,080,365 variant-level associations across the three ancestry groups (**Supplementary Table 3**).

Because not all genes harbored variants from all three functional categories, the effective composition of variant sets in the gene-level analyses varied across genes. As a result, associations identified using different variant sets might reflect the same underlying set of variants (e.g., results obtained using the GOF+LOF+HC-LOF variant set were identical to those obtained using the LOF+HC-LOF variant set for genes that do not harbor any GOF variants). Therefore, to facilitate interpretation of gene-level PheWAS results, we generated effective variant masks representing the variant categories contributing to each test. After processing the summary statistics as described above, we identified a total of 1,856 gene-level and 726 variant-level suggestive associations, as well as 1,204 gene-level significant associations (corresponding to 606 unique gene-trait associations) and 523 variant-level significant associations (corresponding to 430 unique variant-trait associations) across the three ancestry groups.

## Ancestry-specific analyses reveal shared and population-specific well-established associations

To assess shared and unique disease associations of LoF and GoF variants across population groups, we examined the overlap of suggestive PheWAS results among the three ancestry-specific cohorts (**Fig. 3a and 3b**). Notably, the majority of suggestive associations were specific to each population, accounting for 86.8%, 89.7% and 89.5% at the gene-level and 96.6%, 97.6% and 96.5% at the variant-level in the AFR, AMR and EUR populations, respectively. The highest number of unique suggestive associations was observed in the EUR and AFR cohorts, for the gene– and variant-level PheWAS, respectively, probably reflecting larger sample sizes of these two cohorts relative to AMR.

**Fig. 3:**
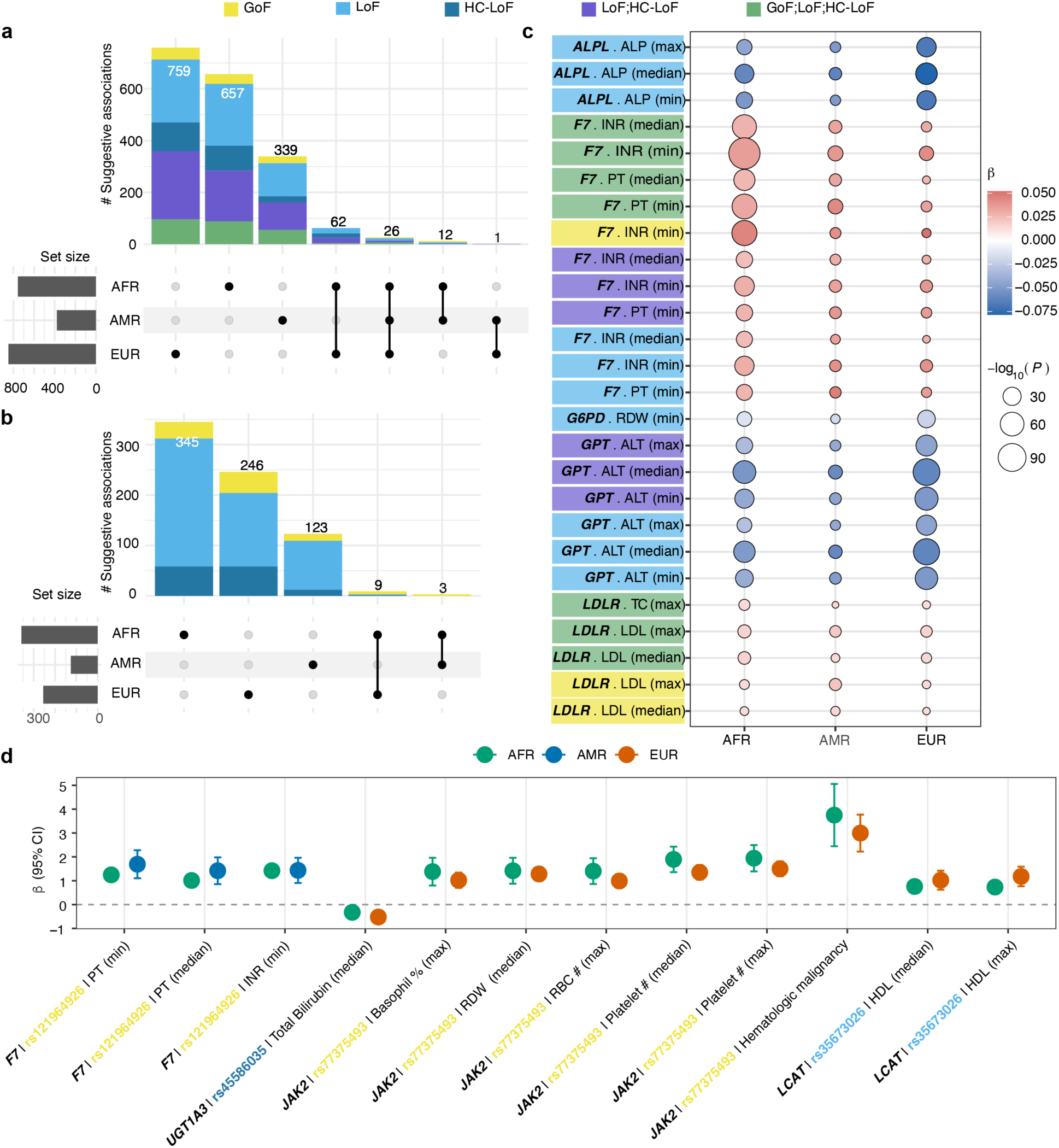
Ancestry-specific suggestive associations. Number of suggestive associations (*P* < 1 x 10^-5^) identified in ancestry-specific **a,** gene-level and **b,** variant-level PheWAS. **c,** Suggestive gene-level associations shared across all three ancestry groups. Dot size indicates the significance level while color indicates the direction (red: increased risk, blue: decreased risk) and magnitude of the effect (darker shade: larger effect sizes). **d,** Suggestive variant-level associations shared between the AFR-AMR and AFR-EUR cohorts. Dots indicate effect size estimates (beta, β) while whiskers indicate 95% confidence intervals for the effect size estimates.

We observed 26 suggestive gene-level associations shared across all three cohorts, including established associations of *ALPL* LoF variants with lower blood alkaline phosphatase levels^15^ (*P_AFR_* = 8.99 x 10^-37^, *P_AMR_* = 2.35 x 10^-12^, and *P_EUR_* = 5.06 x 10^-50^), *F7* GoF, LoF and HC-LoF variants with increased prothrombin time (*P_AFR_* = 5.66 x 10^-68^, *P_AMR_* = 1.33 x 10^-18^, and *P_EUR_* = 3.24 x 10^-09^) and international normalized ratio (INR) levels^16^ (*P_AFR_* = 4.29 x 10^-120^, *P_AMR_* = 4.22 x 10^-19^, and *P_EUR_* = 6.31 x 10^-17^), and *GPT* LoF and HC-LoF variants with lower alanine aminotransferase levels^17^ (*P_AFR_* = 4.40 x 10^-55^, *P_AMR_* = 6.94 x 10^-16^, and *P_EUR_* = 3.16 x 10^-82^) (**Fig. 3c**). On the other hand, variant-level analysis did not reveal any suggestive associations observed in all three cohorts. AFR and EUR shared nine suggestive associations including a *JAK2* GoF variant, rs77375493, with increased risk for hematological malignancies (*P_AFR_* = 1.69 x 10^-08^ and *P_EUR_* = 3.89 x 10^-14^) and increased levels of platelets (*P_AFR_* = 3.59 x 10^-12^ and *P_EUR_* = 4.42 x 10^-18^), red blood cell (RBC) count (*P_AFR_* = 3.77 x 10^-7^ *P_EUR_* = 1.74 x 10^-11^), red cell distribution width (RDW) (*P_AFR_* = 3.49 x 10^-7^ and *P_EUR_* = 2.93 x 10^-17^) and basophil percentage^18^ (*P_AFR_* = 2.99 x 10^-6^ and *P_EUR_* = 4.46 x 10^-10^). Furthermore, AFR and AMR shared three suggestive associations of a *F7* GoF variant, rs121964926, with elevated prothrombin time (*P_AFR_* = 4.06 x 10^-34^ and *P_AMR_* = 1.72 x 10^-8^) and INR levels (*P_AFR_* = 1.40 x 10^-58^ and *P_AMR_* = 9.81 x 10^-8^) (**Fig. 3d**).

Associations that were significant only in the AFR gene-level analysis and not observed as suggestive findings in the other cohorts included those of *GFI1B* LoF and HC-LoF variants with increased mean platelet volume^19^ (*P* = 1.11 x 10^-7^), *ABCA1* LoF and HC-LoF variants with reduced HDL levels^20^ (*P* = 1.14 x 10^-9^), and *ITGB3* GoF variants with reduced platelet counts^21^ (*P* = 9.54 x 10^-7^) (**Supplementary Table 4**). In the AMR-specific analysis, significant findings that were not suggestive in the other cohorts included associations of *PSEN1* LoF and HC-LoF variants with increased risk for dementias and cerebral degeneration^22^ (*P* = 6.35 x 10^-6^), *TSHB* LoF variants with reduced thyroid stimulating hormone levels^23^ (*P* = 1.13 x 10^-6^), and *CP* LoF variants with reduced ceruloplasmin levels^24^ (*P* = 1.32 x 10^-6^) (**Supplementary Table 5)**. For the EUR group, ancestry-specific significant associations included those of *NRXN1* GoF variants with increased risk for psychotic disorders^25^ (*P* = 4.42 x 10^-6^), *SCARB1* LoF variants with elevated high-density lipoprotein (HDL) levels^26^ (*P* = 9.08 x 10^-17^), and *TNFRSF13B* LoF and HC-LoF variants with reduced globulin levels (*P* = 2.70 x 10^-10^), increased albumin-to-globulin ratio (*P* = 1.87x 10^-7^) and reduced monocyte counts^27^ (*P* = 1.46 x 10^-6^) (**Supplementary Table 6**, **Fig. 4a**).

At the variant-level, we replicated well-known associations of rare variants enriched in specific populations, including the associations of *G6PD* LoFs, rs5030872, with Hemoglobin A1c (HbA1c) levels (*P* = 6.17 x 10^-11^) and rs76723693 with lower mean corpuscular volume (*P* = 3.93 x 10^-6^), RDW (*P* = 1.39 x 10^-16^), and HbA1c ^28,29^ (*P* = 7.14 x 10^-11^) in the AFR cohort, *PSEN1* LoF, rs63750082, with increased risk for dementias (*P* = 2.40 x 10^-6^) in the AMR cohort, and *HBB* HC-LoF, rs11549407, with reduced hemoglobin (*P* = 3.91 x 10^-51^), hematocrit (*P* = 4.10 x 10^-26^), and MCH (*P* = 9.78 x 10^-90^) levels and elevated RDW (*P* = 2.07 x 10^-51^), RBC count (*P* = 3.28 x 10^-40^), and total bilirubin levels^30^ (*P* = 9.62 x 10^-6^) in the EUR cohort (**Supplementary Tables 7-9**) (**Fig. 4b**). Overall, these findings replicated previously well-established ancestry-specific and shared associations and provided further evidence related to potential functional mechanisms underlying these associations.

## Multi-ancestry meta-analysis identifies associations not detected in ancestry-specific analyses

We next performed multi-ancestry meta-analyses using traits and genes/variants that were observed in at least two cohorts, corresponding to 51,751,206 and 13,172,910 associations tests at gene– and variant-level, respectively. Of these, 40,550,984 gene-level and 10,675,723 variant-level associations from traits with 0.75 < λ < 1.25 in the synonymous analysis were retained (**Supplementary Table 3**). As a result, we identified 786 gene-level and 317 variant-level associations as significant. Notably, 252 (32.6%) significant gene-level and 224 (70.6%) significant variant-level associations in the multi-ancestry meta-analysis were not observed among suggestive associations in any ancestry-specific analysis. Overall, we identified 364 unique significant gene-trait associations and 267 unique significant variant-trait associations in the multi-ancestry meta-analysis (**Supplementary Tables 10 and 11**).

Significant associations uniquely identified in the gene-level meta-analysis included those of *VNN1* LoF and HC-LoF variants with increased weight (*P* = 7.12 x 10^-8^) and body mass index (BMI) (*P* = 3.34 x 10^-8^) and *ODC1* GoF variants with increased weight (*P* = 8.20 x 10^-9^) (**Fig. 5a**). Both genes have previously been implicated in the regulation of lipogenesis, metabolism, and adiposity^31–33^. Notably, vanin-1 deficiency has been shown to increase adiposity and impair mitochondrial function in brown adipose tissue (BAT), whereas its overexpression protected against diet-induced obesity in mice^31^, consistent with the direction of associations observed in our analysis. Vanin-1 is a glycosylphosphatidylinositol-anchored cell surface protein and can be released into circulation as a soluble form^34^. We therefore performed a focused proteomic evaluation of *VNN1* in a large human cohort, the Multi-Ethnic Study of Atherosclerosis (MESA)^35^, that contained both proteomic and genetic data. Rare LoF and HC-LoF variants of *VNN1* were associated with lower circulating vanin-1 levels (β = –0.74 ± 0.16, *P* = 3.1 x 10^-6^) in MESA (**Fig. 5b**). We separately examined the clinical associations of circulating vanin-1 levels and observed that higher vanin-1 concentrations were associated with higher BMI, weight, and adverse lipid and glycemic profiles (**Fig. 5c and Supplementary Table 12**). Mendelian randomization analyses, however, did not find significant evidence for a causal effect of circulating vanin-1 levels on BMI (β = –0.0006, SE = 0.003, *P* = 0.82) or for whole body fat mass (*β* = –0.003, SE = 0.003, *P* = 0.31). These findings suggest that circulating vanin-1 might partly reflect broader cardiometabolic health and risk rather than specifically mediate the association between *VNN1* loss of function and adiposity. Additionally, tissue-specific effects might not be captured by circulating protein levels, such as those observed in BAT in experimental models.

**Fig. 4:**
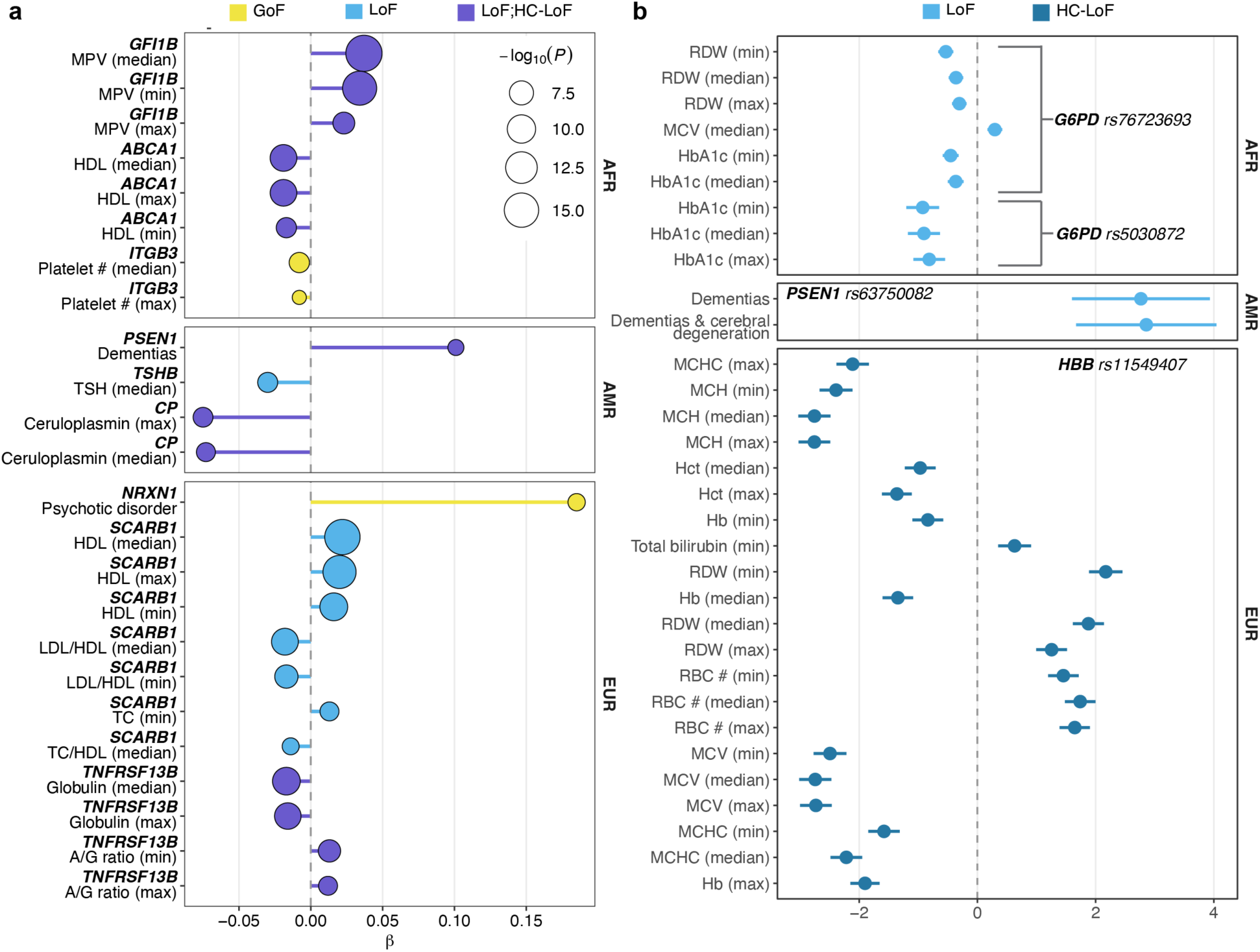
Selected significant associations from ancestry-specific and multi-ancestry meta-analyses. **a**, Examples of ancestry-specific significant associations that were not suggestive in the other ancestry groups at the gene-level. Dots indicate effect size estimates while dot size indicates the significance level **b,** Examples of ancestry-specific significant associations that were not suggestive in the other ancestry groups at the variant-level. Dots indicate effect size estimates while whiskers indicate 95% confidence intervals for the effect size estimates.

**Fig. 5:**
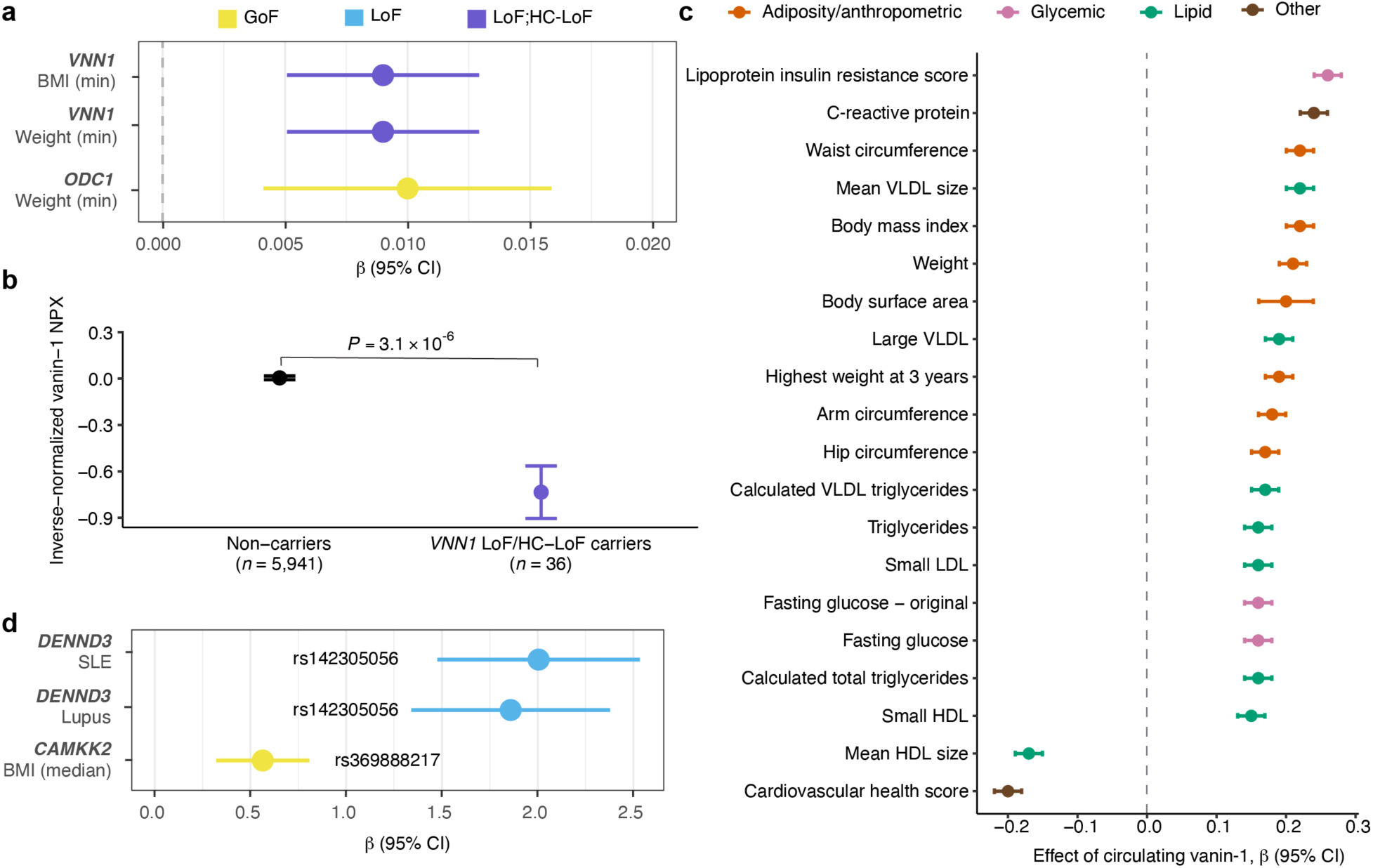
Selected significant associations from multi-ancestry meta-analyses and proteomic analyses of *VNN1*. **a**, Examples of significant associations identified in the multi-ancestry meta-PheWAS that were not suggestive in the ancestry-specific analyses at the gene-level. **b,** Circulating vanin-1 levels in carriers and non-carriers of rare *VNN1* LoF and HC-LoF variants. NPX: normalized protein expression. **c,** Top 20 cardiometabolic traits associated with circulating vanin-1 levels. **d,** Examples of significant associations identified in the multi-ancestry meta-PheWAS that were not suggestive in the ancestry-specific analyses at the variant-level. Dots indicate effect size estimates while whiskers indicate 95% confidence intervals for the effect size estimates.

At the variant-level, rs142305056, a predicted LoF variant in *DENND3* was associated with an increased risk for systemic lupus erythematosus (*P* = 1.93 x 10^-7^), consistent with the role of *DENND3* in promoting autophagy and supporting the previous evidence implicating this gene in the regulation of inflammation^36,37^. Moreover, rs369888217, a predicted GoF variant in *CAMKK2*, was associated with increased BMI (*P* = 5.48 x 10^-6^) (**Fig. 5d**). Previous studies have shown that *CAMKK2* is a key regulator of energy homeostasis and metabolism by stimulating appetite and promoting glucose production, and that deficiency of *CAMKK2* protects against high fat diet induced obesity in mice^38,39^. Notably, neither of these associations was identified in the ancestry-specific PheWAS. Together, these findings suggest the enhanced power of meta-analysis for capturing significant associations that might remain undetected in ancestry-specific analyses and emphasizes the importance of performing both ancestry-specific and multi-population analyses.

## Identification of known and previously unreported findings and replication analyses in UK Biobank and All of Us Research Program

Across the ancestry-specific and multi-ancestry meta-analyses, we identified numerous well-established associations as well as previously unreported findings that are not represented in ClinVar^40^, Human Gene Mutation Database (HGMD) Professional version^41^ and/or the NHGRI-EBI GWAS Catalog^42^ based on our predefined criteria (see Methods). Of the 630 unique gene-trait associations, 436 (69.2%) have not been reported in ClinVar/HGMD or the GWAS catalog (**Supplementary Tables 4-6 and 10**). Similarly, 388 (61.1%) of the 635 unique variant-trait associations have not been reported in these resources (**Supplementary Tables 7-9 and 11**). Although the proportions of previously unreported significant associations were comparable for the three ancestry groups, the highest proportions were observed in the AMR cohort, followed by the AFR and EUR cohorts in gene– and variant-level analyses in both ClinVar/HGMD and GWAS catalog (**Extended Data Fig. 5a and 5b**). Across the variant categories, LoF variants exhibited the highest proportions of previously unreported significant associations in both gene-level and variant-level analyses (**Extended Data Fig. 5c and 5d**).

We then evaluated the replication of the gene-level significant associations from the MSM EUR-specific analysis in the UK Biobank^43^, a cohort composed predominantly of individuals of EUR ancestry, using cardiovascular and endocrine/metabolic binary traits, as well as cardiometabolic quantitative traits, such as blood pressure, glucose, and cholesterol measurements (**Supplementary Table 13**). Of the 127 evaluable associations, 83 (65.4%) have been reported in ClinVar/HGMD and/or the GWAS Catalog, of which 78 (93.9%) displayed a consistent direction of effect and 72 (86.8%) replicated with a *P* < 0.05. Among the 44 previously unreported associations, 24 (54.5%) showed a consistent direction of effect, and two replicated at nominal significance level (**Fig. 6a**). One of the replicated associations was that of *NOX5* HC-LoF variants with an increased risk of ventricular premature depolarization (*P_EUR_* = 2.73 x 10^-6^ and *P_UKBB_* = 2.15 x 10^-2^). *NOX5* is involved in regulation of oxidative stress and has been implicated in activation of MAPK-mediated cardiac remodeling^44^. The other replicated association was between *XXYLT1* LoF variants and an increased risk of disorders of lipoprotein metabolism and other lipidemias (*P_EUR_* = 2.25 x 10^-6^ and *P_UKBB_* = 3.43 x 10^-3^). *XXYLT1* encodes a xyloside xylosyltransferase involved in protein O-linked glycosylation and has previously been implicated in venous thromboembolism through shared genetic regulation in adipose tissue and arterial tissues^45^. However, its role in lipoprotein metabolism remains to be determined.

**Fig. 6:**
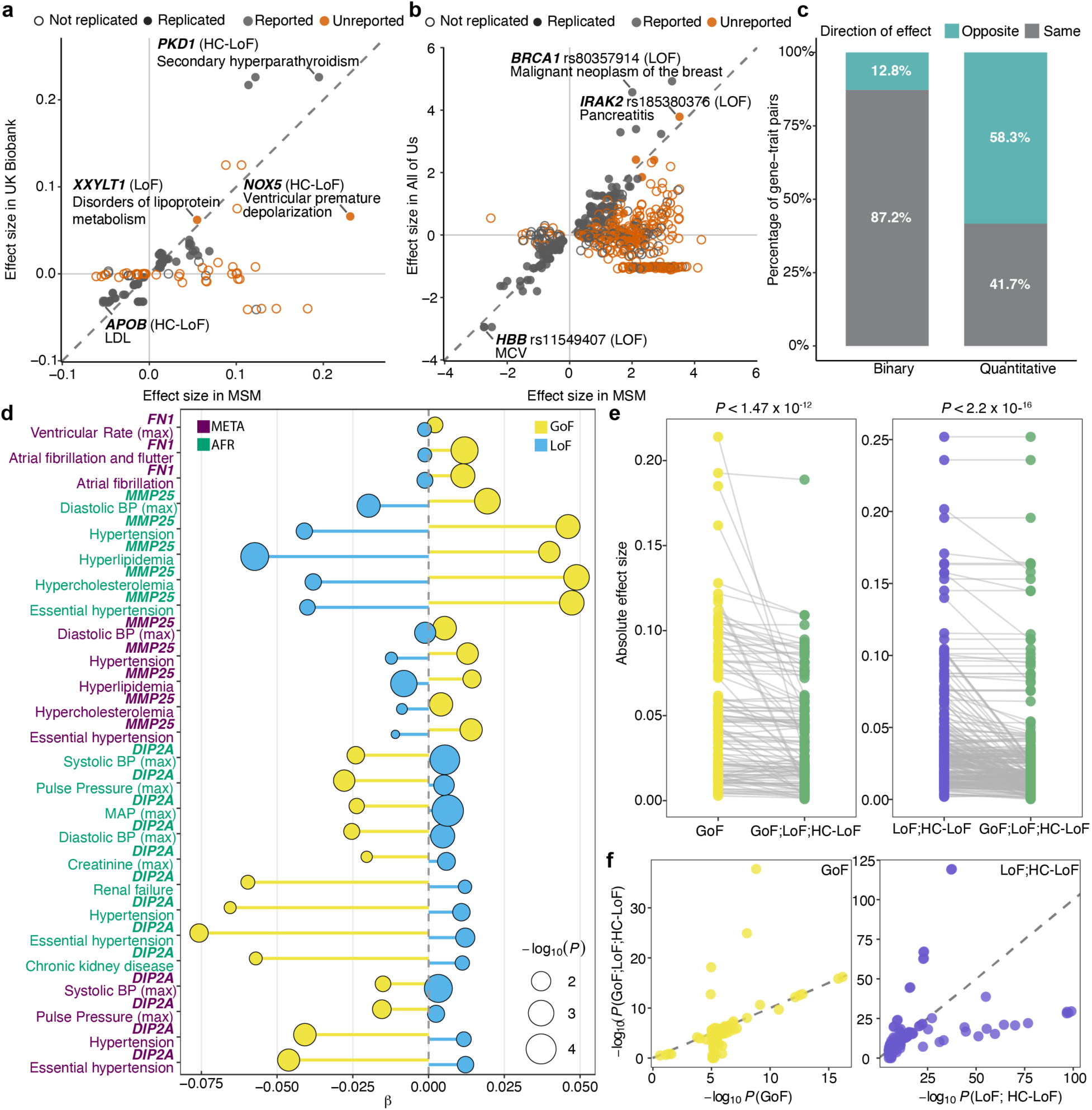
Replication analyses and assessment of the direction of effect. **a**, Distribution of effect sizes for gene-level significant cardiometabolic associations in the MSM (discovery cohort) and the UK Biobank (replication cohort), (*n* = 127 associations). **b,** Distribution of effect sizes for variant-level significant associations in the MSM (discovery cohort) and the AoU (replication cohort) (*n* = 563 associations). Six associations with effect sizes > 4 are not shown (see Supplementary Table 14). **c,** Proportion of associations exhibiting opposite directions of effect in the GoF– and LoF-stratified analyses. **d,** Examples of associations exhibiting opposite directions of effect in the GoF– and LoF-stratified analyses. **e,** Comparison of effect sizes from the combined analysis with those from the functionally stratified analyses. **f,** Comparison of *P* values from the combined analysis with those from the functionally stratified analyses. For e, and f, statistical significance was assessed using one-sided paired Wilcoxon signed ranks tests (n = 112 and 237 associations for GoF versus combined and LoF;HC-LoF versus combined analyses, respectively).

For the replication of variant-level significant findings, we used ancestry-specific and meta-analysis summary statistics from All of Us (AoU)^9^ “All by All” release (**Supplementary Table 14**). Of the 569 evaluable associations, 280 (49.2%) have been previously reported in ClinVar/HGMD and/or the GWAS Catalog, of which 221 (78.9%) showed a consistent direction effect and 158 (71.5%) replicated with a *P* < 0.05. Among the 289 previously unreported associations, 121 (41.9%) displayed a consistent direction of effect and five replicated at nominal significance (**Fig. 6b**). These included the association of an *IRAK2* LoF variant, rs185380376, with an increased risk of pancreatitis in the AMR cohort (P*_AMR_* = 2.04 x 10^-6^ and *P_AoU-AMR_* = 8.94 x 10^-3^). *IRAK2* encodes the interleukin-1 receptor-associated kinase 2, a key regulator of innate immunity, and its deficiency has been linked to immune dysregulation^46^. Given that innate immunity is an integral part of the pathogenesis of pancreatitis^47,48^, this association warrants further investigation. Overall, replication analyses demonstrated substantial concordance for previously reported associations and identified several previously unreported associations with supporting findings across independent datasets.

## Assessment of opposite directions of effect of GoF and LoF variants on the same phenotypes

To evaluate whether GoF and LoF variants exhibited different directions of effect across associated phenotypes in the 3,680 genes harboring both variant categories, we examined the association results of these genes in ancestry-specific and multi-ancestry meta-analyses. Because the number of significant or suggestive gene-trait pairs tested with both GoF and LoF variant sets was limited, we extended the assessment to include associations at nominal significance (*P* < 0.05). Among 5,666 unique gene-binary trait pairs for which both GoF-only and LoF-only analyses were nominally significant, 723 (12.7%) displayed opposite directions of effect between the two variant categories. In contrast, of the 1,690 unique gene-quantitative trait pairs, 985 (58.3%) showed opposite directions of effect, suggesting that opposing effects of gene function are more frequently observed for quantitative traits compared to binary phenotypes (**Fig. 6c**).

Notable examples showing consistent opposite directions of effect across both binary and quantitative traits in the multi-ancestry meta-analysis included *DIP2A*, *FN1* and *MMP25* genes (**Fig. 6d and Supplementary Table 15**). *DIP2A* GoF variants were associated with a reduced risk for the diagnosis of hypertension and essential hypertension, as well as lower systolic blood pressure and pulse pressure measurements, whereas LoF variants were associated with an increased risk of these conditions and higher values of blood pressure measurements. In the AFR-specific analysis, the same associations were observed, with additional opposing effects of GoF and LoF variants on chronic kidney disease and renal failure, as well as mean arterial pressure and blood creatinine levels. Although *DIP2A* has not previously been implicated in essential hypertension, its cardiovascular protective role as an endothelial follistatin-like 1 (FSTL1) receptor provides biological plausibility for the observed blood pressure and kidney-related associations^49,50^.

Different directions of effect were also observed for *FN1,* which encodes fibronectin. GoF variants were associated with an increased risk for the diagnosis of atrial fibrillation and flutter, as well as higher ventricular rate measured by electrocardiography, whereas LoF variants displayed the opposite pattern. Fibronectin is a major extracellular protein involved in cardiac extracellular remodeling and fibrosis, which are important processes underlying atrial fibrillation^51^. Lastly, *MMP25* GoF variants were associated with an increased risk for the diagnosis of hypertension, essential hypertension, hyperlipidemia, and hypercholesterolemia, as well as higher values of diastolic blood pressure, while LoF variants exhibited the opposite direction of effect. *MMP25*, a member of the matrix metalloproteinase family, has recently been implicated in pathological vascular remodeling and atherosclerosis in experimental models^52^. These findings support the biological relevance of distinguishing GoF and LoF variants as they might yield distinct phenotypic effects and suggest that combining them into a single variant set in gene-based analyses can obscure association signals.

## Collapsing predicted GoF and LoF variants together attenuates gene-based association signals

To evaluate the impact of collapsing variants with different predicted functional consequences, we compared the association results from genes harboring both GoF and LoF/HC-LoF variants. We focused on significant associations identified in analyses with GoF-only and LoF+ HC-LoF effective masks and compared them with those from the combined mask (GoF+LoF+HC-LoF), which represents variant sets typically obtained from general pathogenicity predictors that do not distinguish functional effects.

Across analyses of different ancestry groups and multi-ancestry meta-analysis, collapsing variants with different predicted functional impact resulted in smaller effect sizes for 80.4% of significant GoF associations and 78.9 % significant LoF+HC-LoF associations with median attenuations of 10.3% and 17.0% respectively (One-sided paired Wilcoxon signed-rank test *P* < 2.2 x 10^-16^ and *P* = 1.47 x 10^-12^) (**Fig. 6e**). The significance of associations was also reduced for 58.4% GoF and 62.0% LoF+HC-LoF associations (One-sided paired Wilcoxon signed-rank test, GoF, *P* = 2.64 x 10^-7^ and LoF+HC-LoF, *P* = 2.57 x 10^-6^, respectively) (**Supplementary Table 16**). Moreover, 43.8% of GoF and 18.9% LoF+HC-LoF significant associations from the functionally stratified analysis were not even suggestive in the combined analysis. Collectively, these findings demonstrate that collapsing predicted GoF and LoF variants together can dilute association signals by reducing statistical significance and/or effect size estimates, emphasizing the importance of distinguishing different functional consequences in gene-based tests to improve detection of gene-trait associations.

## Discussion

In this study, we conducted a systematic phenome-wide analysis of rare predicted GoF and LoF variants across 18,214 genes and 1,446 traits in a genetically diverse biobank comprised of 50,067 participants. By stratified analysis of GoF, LoF and HC-LoF variants, we identified numerous established and previously unreported gene– and variant-level associations, providing further insight into potential functional mechanisms of the identified associations. High genetic diversity of the MSM cohort enabled identification of both ancestry-specific and shared associations. Although the majority of suggestive associations were specific to each ancestral group, several well-established associations were consistently observed across the three groups, supporting the robustness of our analytical framework.

Numerous ancestry-specific findings reflected the effects of rare variants observed predominantly in specific populations. Accordingly, our study in ancestrally and phenotypically diverse MSM makes a unique contribution to large-scale PheWAS efforts, which have mostly included individuals of European ancestry^5,6^ or focused on common variant associations^7^, by also providing functionally stratified association results. The higher prevalence of many phenotypes in the AFR and AMR cohorts further emphasizes the importance of including ancestrally diverse populations in rare variant association studies, although these differences likely reflect not only underlying disease burden but also varying levels of healthcare utilization and clinical ascertainment.

Our results showed that predicted GoF and LoF variants frequently exhibit different phenotypic effects. Opposite effects were more commonly observed in quantitative traits, which might reflect more direct consequences of altered gene function than disease diagnoses that are influenced by heterogenous disease mechanisms. Associations of predicted GoF and LoF variants in *DIP2A*, *FN1* and *MMP25* exemplify how altered gene function in opposite directions can produce opposing effects across both laboratory measurements and disease phenotypes. Such bidirectional effects also have important implications for therapeutic target discovery and can provide insight into expected consequences of pharmacological activation or inhibition. Accordingly, human genetic evidence and the direction of genetic effects inferred by LoGoFunc have been instrumental in the prioritization of therapeutic targets^53,54^. However, GoF and LoF variants in the same gene can also be associated with the same phenotype, as observed for *F7*, where both predicted GoF and LoF variants were associated with increased PT and INR. These associations are not necessarily contradictory. LoF variants in *F7* are well established to cause factor VII deficiency and consequently prolong PT^16^. On the other hand, some F7 GoF variants with increased FVII activity in vitro cause paradoxically prolonged PT in humans, potentially due to reduced secretion of altered FVII into the circulation^55^. This example demonstrates how GoF and LoF definitions might depend on the context in which the variant function is assessed, including effects on protein activity, expression, cellular pathways, tissues and organism-level phenotypes. The training data used for LoGoFunc largely reflected the GoF and LoF definitions based on expression and protein-level functional effects, consistent with definitions mostly used in previous studies^4^. Therefore, predicted GoF and LoF classifications and their associations should be interpreted in the context of these definitions, as the direction of downstream effects might not be uniform across different biological levels.

We also showed that stratifying variants according to predicted functional effects in gene-level analyses might provide stronger association signals than the conventional approach of aggregating variants irrespective of their functional consequences. Most current gene-based association studies aggregate variants based primarily on predicted pathogenicity or protein consequence without explicitly considering whether variants are expected to increase or decrease protein function. However, variants with opposing biological effects might partially counteract in gene-based collapsing tests, reducing statistical power and obscuring association signals^56^. Consistently, we observed attenuation of both effect sizes and statistical significance after collapsing predicted GoF and LoF variants into a single variant set, while a considerable proportion of significant associations identified using functionally stratified analyses were no longer detected in the combined analyses. These findings suggest that incorporating predicted molecular mechanisms can improve the performance of rare variant association studies beyond conventional pathogenicity-based variant aggregation.

More than half of the significant associations identified in this study have not previously been reported in ClinVar, HGMD, or the GWAS Catalog. Although additional validation will be required to establish the biological relevance of these findings, the large number of previously unreported associations likely reflects several distinguishing features of our study, including functional stratification of predicted GoF and LoF variants, analysis of a large array of quantitative clinical traits along with binary diagnoses, and the inclusion of genetically diverse populations. Replication analyses demonstrated high concordance for previously established associations and provided supporting evidence for several previously unreported findings, although replication rates were lower for novel associations. This is expected for rare variants, many of which exhibit substantial population specificity and are challenging to replicate across cohorts with different ancestry compositions, phenotype definitions, sequencing platforms, and healthcare systems.

There are several limitations to our study. First, GoF and LoF classifications were based on computational predictions and misclassifications might occur as illustrated by the observed association of *LDLR* GOF variants with elevated LDL levels. Therefore, definitive determination of GoF and LoF effects requires experimental validation. Although the improved association signals support the utility of these predictions, variant function is often context dependent and might not be fully captured by current prediction tools trained on datasets that use varying definitions of GoF and LoF. Second, replication of previously unreported associations was limited, likely reflecting the inherent challenges of studying rare variants across different cohorts with varying ancestry compositions and phenotype definitions^57^. Lastly, despite restricting our analyses to rare variants, some of the observed associations might reflect linkage disequilibrium with nearby causal variants in other genes rather than direct effects of the identified variants or genes themselves^58^, exemplified by the observed association of *OR52R1* LoF variants with hemoglobinopathy-related traits, which likely arises from proximity to *HBB*.

In conclusion, our findings demonstrate the importance of incorporating predicted GoF and LoF effects into rare variant association studies and establish a comprehensive phenome-wide resource linking predicted GoF and LoF variation to human disease. For rapid exploration of GoF and LoF associations, we developed a publicly available web browser (https://goflofphewas.mssm.edu) that provides access to ancestry-specific and multi-ancestry gene– and variant-level PheWAS summary statistics. This work and future efforts toward functionally informed analysis of rare coding variation can provide insights into disease mechanisms and facilitate therapeutic target discovery.

## Methods

### Mount Sinai Million Health Discoveries Program

The MSM is a hospital-based biobank within the New York City health system, launched in 2022 with the goal to integrate genetics into real-world clinical care by leveraging genetic and clinical data from a highly diverse patient population (https://icahn.mssm.edu/research/ipm/programs/mount-sinai-million). Participants were recruited from primary care clinics across the Mount Sinai Health System without specific inclusion criteria, thus providing a cohort broadly representative of the New York City population and its neighborhood.

### Whole exome sequencing and quality control

Exome sequencing was performed by Regeneron (NY, USA) using the IDT xGen (*n* = 31,685) and TWIST (*n* = 30,215) capture kits on an Illumina HiSeq 2500 platform. Raw reads were aligned to the GRCh38 reference genome using Burrows-Wheeler Aligner (BWA)^59^. Sample-level genomic variant calling format (gVCF) files were generated using DeepVariant and joint genotyping was performed using GLnexus v1.4.5^60^.

Samples with discordant reported sex and genetically inferred sex, duplicate samples, as well as those with an exome coverage < 80% at 20X or contamination > 5% were removed from the dataset. The mean proportion of target bases covered at ≥ 20X was 94.8% across the remaining samples. Variants that fell into the intersection of the two capture kits were extracted and those with an average genotype quality (GQ) < 20, average depth (DP) <10, a phred-scaled quality score (QUAL) < 20, an allele balance < 0.2, a missingness >10% or a deviation from Hardy-Weinberg equilibrium with a *P* < 1×10⁻¹⁵ were excluded. Normalization of insertion-deletion variants (indel), splitting of multi-allelic variants, and calculation of QC statistics were carried out using BCFtools^61^. After the quality control (QC) steps, the cohort included 58,990 samples and 5,799,823 variants.

### Genetic ancestry inference

Genetic ancestries of MSM participants were inferred using HapMap3 populations as a reference^10^. Principal components (PCs) were calculated from HapMap3 samples using common SNPs with a minor allele frequency (MAF) > 5%, and MSM samples were projected onto these PCs. Kernel density estimates (KDEs) were then trained for each ancestral group using the first four PCs. Based on the KDE results, each MSM sample was assigned to one of five superpopulations according to its genetic similarity to the HapMap3 reference populations: African (AFR, *n* = 20,209), Admixed American (AMR, *n* = 8,657), East Asian (EAS, *n* = 2,504), European (EUR, *n* = 22,065), South Asian (SAS, *n* = 2,138) or unknown (*n* = 3,417). Due to limited sample sizes of the EAS and SAS groups, the downstream analyses were restricted to the 50,067 samples from the AFR, AMR, and EUR groups.

### Constructing the MSM phenome

Phenotyping was performed using EHR data from MSM participants. Binary traits were defined by mapping ICD-9 and ICD-10 codes to 3,200 clinically meaningful phenotype codes (phecodes) using phecodeX v.1.0^11^. For each ancestral group, binary phenotypes with at least 50 cases were selected. Quantitative traits were manually curated from laboratory and body measurements of MSM participants, and traits observed in at least 50 individuals and passing the QC were retained in each ancestral group. For each measurement, the median, minimum, and maximum values were calculated per participant. Extreme outliers, defined as values more than ± 6 standard deviation from the trait mean, were excluded for each quantitative trait. Participants without linked EHR data were also excluded. The final dataset included 19,856 AFR, 8,531 AMR, and 21,680 EUR participants.

For the comparison of disease prevalence across phecode categories, age– and sex-standardized prevalence estimates were obtained using the direct standardization method implemented in the PHEindicatormethods R package version 2.1.1^62^. The age and sex distribution of the combined study population was used as the standard population.

### Identification of predicted LoF and GoF variants

WES data from 50,067 participants that include 4,900,775 high-quality variants were annotated using the Ensembl Variant Effect Predictor (VEP) v112^63^. Those with a predicted “HIGH” impact by VEP were further annotated using LOFTEE^13^, and only high confidence LoFs were retained. In addition, missense variants predicted as either LoF or GoF by LoGoFunc^4^ were selected. Then pLoF and pGoF variants with a POPMAX MAF less than 1% in gnomAD v4.0.1^12^ exomes and genomes were retained. In total, 596,454 variants in 18,214 genes were included in downstream analyses. For the synonymous model, 1,808,714 synonymous variants in the same genes were selected using the same gnomAD MAF threshold.

### PheWAS

The Scalable and Accurate Implementation of Generalized Mixed Model (SAIGE)^14^ was employed to conduct ancestry-specific PheWAS in each ancestral group. First, a sparse genetic relationship matrix (GRM) was constructed using 5,000 linkage disequilibrium (LD)-pruned random markers with a relatedness cut-off of 0.125 for each ancestral group. In the step 1 of SAIGE, 2,000 random markers were selected based on minor allele count (MAC) thresholds: 1,000 markers with 10 ≤ MAC < 20 and 1,000 random markers with MAC ≥ 20. Null logistic mixed models for binary traits and null linear mixed models for inverse-normalized quantitative traits were fitted for each ancestral group by adjusting for age, sex, batch and first ten PCs using the sparse GRMs. In step 2 of SAIGE, set-based tests using variants with a MAF < 1% and single-variant tests using variants with a MAC ≥ 10 were conducted using five variant masks: GoF, LoF, HC-LoF, LoF:HC-LoF, and GoF:LoF:HC-LoF, as well as using synonymous variants.

A fixed-effect multi-ancestry meta-analysis of the ancestry-specific variant-level PheWAS results was performed using GWAMA^64^ by adjusting for genomic control. Gene-level meta-analysis was conducted using aggregated Cauchy association test (ACAT) method implemented in metafor package^65^ on gene-level ancestry-specific PheWAS results. SKAT-O *P* values from ancestry-specific PheWAS results were combined using ACAT by weighting the results according to sample size. In addition, *P* and beta values derived from burden test results from SAIGE were meta-analyzed using an inverse variance weighting (IVW) method to obtain meta-analysis burden *P* and beta values. Heterogeneity between AFR, AMR and EUR cohorts was assessed based on direction of effect in each population as well as Cochran’s heterogeneity statistic (Q) and heterogeneity index (I^2^). Associations observed in at least two cohorts were included for both variant– and gene-level meta-analyses.

### Quality control of the summary statistics

Because PheWAS inherently includes a large number of correlated traits with highly varying case numbers and the primary objective of the study was identifying genes associated with each trait for a given functional variant class, multiple testing correction was performed using the BH-FDR method separately for each combination of trait and variant set rather than calculating a global FDR. The same strategy was applied to the variant-level PheWAS for each combination of trait and variant category. Adjusted *P* values < 0.05 were considered statistically significant.

To obtain high-quality association results, we removed the tests with a cumulative trait MAC < 10 in gene-level analyses and a trait MAC < 10 in variant-level analyses, based on improved calibration observed in the synonymous variant analysis. We then recalculated λ values for each trait and excluded those with a synonymous lambda < 0.75 and > 1.25.

### Identification of known and previously unreported associations

We used ClinVar^40^ (accessed on 02/18/2026), HGMD professional version 2026.2^41^ and the NHGRI-EBI GWAS Catalog^42^ to identify the significant gene-trait and variant-trait associations that had been previously reported. Phenotype matching between the study traits and the three databases was performed using SapBERT-assisted semantic matching^66^, followed by manual review of all candidate matches to identify clinically or biologically equivalent or closely related phenotypes. For gene-level analyses, an association was considered previously reported if the corresponding gene had been linked to a relevant phenotype through a pathogenic or likely pathogenic variant in ClinVar, a non-retracted disease associated variant in HGMD or as a mapped gene in the GWAS catalog. For variant-level analyses, an association was considered previously reported if the exact variant was reported for a relevant phenotype in ClinVar or HGMD. For GWAS Catalog comparisons of variant-level findings, GWAS Catalog associations within ± 500 kb of the identified variant were examined.

### *VNN1* proteomic analysis

The Multi-Ethnic Study of Atherosclerosis (MESA)^35^ is a longitudinal population-based adult cohort that aimed to study the epidemiology of subclinical cardiovascular disease. In brief, 6,814 participants that were free of overt cardiovascular disease (aged 45-84 years old that were self-reported as White, Black, Hispanic, or Chinese) were recruited and enrolled across the U.S. with exam 1 occurring from 2000-2002. Olink proteomics was conducted using Proximity Extension Assay (PEA) technology to measure 2,943 circulating proteins including vanin-1 on fasting plasma samples obtained at exam 1 that were stored at –80°C. In brief, samples were then thawed, serially diluted based on platform recommendations, and underwent a dual-recognition immunoassay that results in the hybridization and extension of complementary oligonucleotides bound to the antibody pair targeting vanin-1. The resulting amplicon is then detected and quantified using next-generation sequencing. Measurements were reported in relative quantification units (Normalized Protein Expression, NPX) on a log2 scale. The mean intra-plate coefficient of variation (CV) for vanin-1 was 3.4%. Vanin-1 associations with cardiometabolic traits were modeled using linear regression adjusted for age, sex, race and ethnicity, and batch for all continuous clinical variables that were also measured at MESA exam 1. The clinical variable was the outcome and Vanin-1 NPX values were the dependent variable and were standardized with mean = 0 and SD = 1.

Whole genome sequencing was completed in MESA through the National Heart, Lung, and Blood Institute’s (NHLBI) Trans-Omics for Precision Medicine (TOPMed) program^67^ with detailed methods reported https://topmed.nhlbi.nih.gov/data-resources/methods. 12 LoF and 2 HC-LoF variants with a gnomAD v4.1 POPMAX MAF less than 1% were identified in *VNN1*. A gene-based collapsing test was performed to examine the effect of rare LoF and HC-LoF variants in *VNN1* on inverse rank normalized vanin-1 NPX using linear regression adjusted for age, sex, and the first 10 principal components.

We conducted two-sample cis-Mendelian Randomization (MR) to determine if Vanin-1 circulating levels were potentially causally associated with select cardiometabolic traits. We identified cis-region single-nucleotide variants (SNVs) associated with circulating protein concentrations (i.e., protein quantitative trait loci, pQTLs) utilizing a GWAS of Olink protein from the UK Biobank^68^. A cis variant was defined as within 1 Mb of the protein gene-coding region. Risk alleles were modeled as having an additive effect in logistic regression models adjusted for age, biological sex, and the first 10 genetic principal components. Independent SNVs were defined using a LD threshold of *r*^2^ < 0.01 and *P* < 5×10^-5^. SNVs with F-statistics < 10 was used to exclude weak protein instruments. Data from previously published GWAS for BMI^69^ and whole-body fat mass^70^ were used for the outcome. Allelic effects were harmonized. The effect of genetically quantified protein concentrations on the individual cardiometabolic outcomes was assessed with IVW methods. Steiger filtering was used to control for reverse causation. FDR significance was used, correcting for the total number of *cis* variants tested in each MR method. The R statistical package TwoSampleMR (v0.7.4)^70^ was used.

### Replication analyses of gene– and variant-level findings

For the replication analyses of the gene-level findings, we evaluated the significant associations identified in the MSM EUR analysis. We used the phecodes from the cardiovascular and endocrine/metabolic categories, as well as lab and vital measures related to cardiometabolic traits, namely total cholesterol, LDL cholesterol, HDL cholesterol, total cholesterol/HDL ratio, LDL/HDL ratio, triglycerides, hemoglobin A1c, blood glucose systolic and diastolic blood pressure, and mean arterial pressure.

Replication analyses were performed using the UK Biobank 200k WES dataset^43^. Genetic ancestries of UK Biobank participants were inferred using PCA and ADMIXTURE^71^, and 180,500 EUR samples were included in the analyses. ICD-9 and ICD-10 codes were mapped to phecodeX definitions, and the exact phecode matching was performed when possible. If the corresponding phecode had fewer than 50 cases in the UK Biobank, the parent phecode was used instead. For quantitative traits with two measurements per participant, the average value was calculated. Per sample averages of the quantitative traits were calculated when there were multiple such measurements in UK Biobank. Rare GoF, LoF and HC-LoF variants were identified, and gene-level association testing was performed using SAIGE by following the same methodology as in the MSM analysis by including, age, sex, and first ten PCs as covariates.

The All of Us Research Program Controlled Tier Dataset v8^9^ was used for the replication analyses of the variant-level significant associations in MSM. *All by All* summary statistics were accessed through the All of Us Researcher Workbench. Variant-level association results corresponding to the significant associations identified in each ancestry-specific and multi-ancestry meta-analysis were extracted. For binary traits, the exact phecode matching was performed whenever possible, otherwise the corresponding parent phecode was used. For the quantitative traits, association results of the same measurement or the closest available proxy (e.g., INR and PT) were used.

## Data availability

PheWAS summary statistics are publicly available at our web browser https://goflofphewas.mssm.edu and at https://zenodo.org/records/22770086^72^. Functional predictions generated with LoGoFunc are available at https://itanlab.shinyapps.io/goflof/ and https://zenodo.org/records/10126185^73^. Mount Sinai Million Health Discoveries Program: https://mountsinaimillion.org/en/contact

## Code availability

Code to train and test LoGoFunc is available at https://gitlab.com/itan-lab/logofunc and https://zenodo.org/record/7916161^74^.

## Acknowledgments

We gratefully acknowledge Mount Sinai Million Health Discoveries Program, UK Biobank, All of Us Research Program and MESA participants for their contributions, without whom this research would not have been possible. The Mount Sinai Million Health Discoveries Program data used in the study was accessed with approval from the Institutional Review Board at the Icahn School of Medicine at Mount Sinai (STUDY-20-00338), and all participants provided informed consent. The use of data from the UK Biobank was approved with the UK Biobank Resource under application number 53074. We also thank the National Institutes of Health’s All of Us Research Program for making available the data included in replication analyses in this study.

For the Multi-Ethnic Study of Atherosclerosis (MESA), whole genome sequencing (WGS) for the Trans-Omics in Precision Medicine (TOPMed) program was supported by the National Heart, Lung and Blood Institute (NHLBI). WGS for “NHLBI TOPMed: Multi-Ethnic Study of Atherosclerosis (MESA)” (phs001416.v1.p1) was performed at the Broad Institute of MIT and Harvard (3U54HG003067-13S1). Centralized read mapping and genotype calling, along with variant quality metrics and filtering were provided by the TOPMed Informatics Research Center (3R01HL-117626-02S1). Phenotype harmonization, data management, sample-identity QC, and general study coordination, were provided by the TOPMed Data Coordinating Center (3R01HL-120393-02S1). The Multi-Ethnic Study of Atherosclerosis (MESA) projects are conducted and supported by the National Heart, Lung, and Blood Institute (NHLBI) in collaboration with MESA investigators. Support for MESA is provided by contracts 75N92025D00022, 75N92020D00001, HHSN268201500003I, N01-HC-95159, 75N92025D00026, 75N92020D00005, N01-HC-95160, 75N92020D00002, N01-HC-95161, 75N92025D00024, 75N92020D00003, N01-HC-95162, 75N92025D00027, 75N92020D00006, N01-HC-95163, 75N92025D00025, 75N92020D00004, N01-HC-95164, 75N92025D00028, 75N92020D00007, N01-HC-95165, N01-HC-95166, N01-HC-95167, N01-HC-95168, N01-HC-95169, UL1-TR-000040, UL1-TR-001079, UL1-TR-001420, UL1TR001881, and R01HL105756. The authors thank the MESA investigators and staff for their valuable contributions. A full list of participating MESA investigators and institutions can be found at http://www.mesa-nhlbi.org.

## Author contributions

**M.E.K.**: Conceptualization, Data curation, Formal analysis, Methodology, Writing – original draft, Writing – review & editing. **D.S.**: Software. **Y.W.**: Data curation. **Z.C.**: Formal analysis, Writing – review & editing **S.D.**: Formal analysis. **H.A.**: Writing – review & editing. **P.D.S.**: Resources. **D.N.C.**: Resources. **L.M.R.**: Resources. **L.A.L.**: Resources. **M.T.**: Resources. **M.O.G.**: Resources. **S.S.R.**: Resources. **J.I.R.**: Resources. **P.C.**: Writing – review & editing. **R.E.G.**: Resources, Writing – review & editing. **Y.I.**: Conceptualization, Supervision, Writing – review & editing. All authors reviewed and approved the final version of the manuscript.

**Extended Data Fig. 1:**
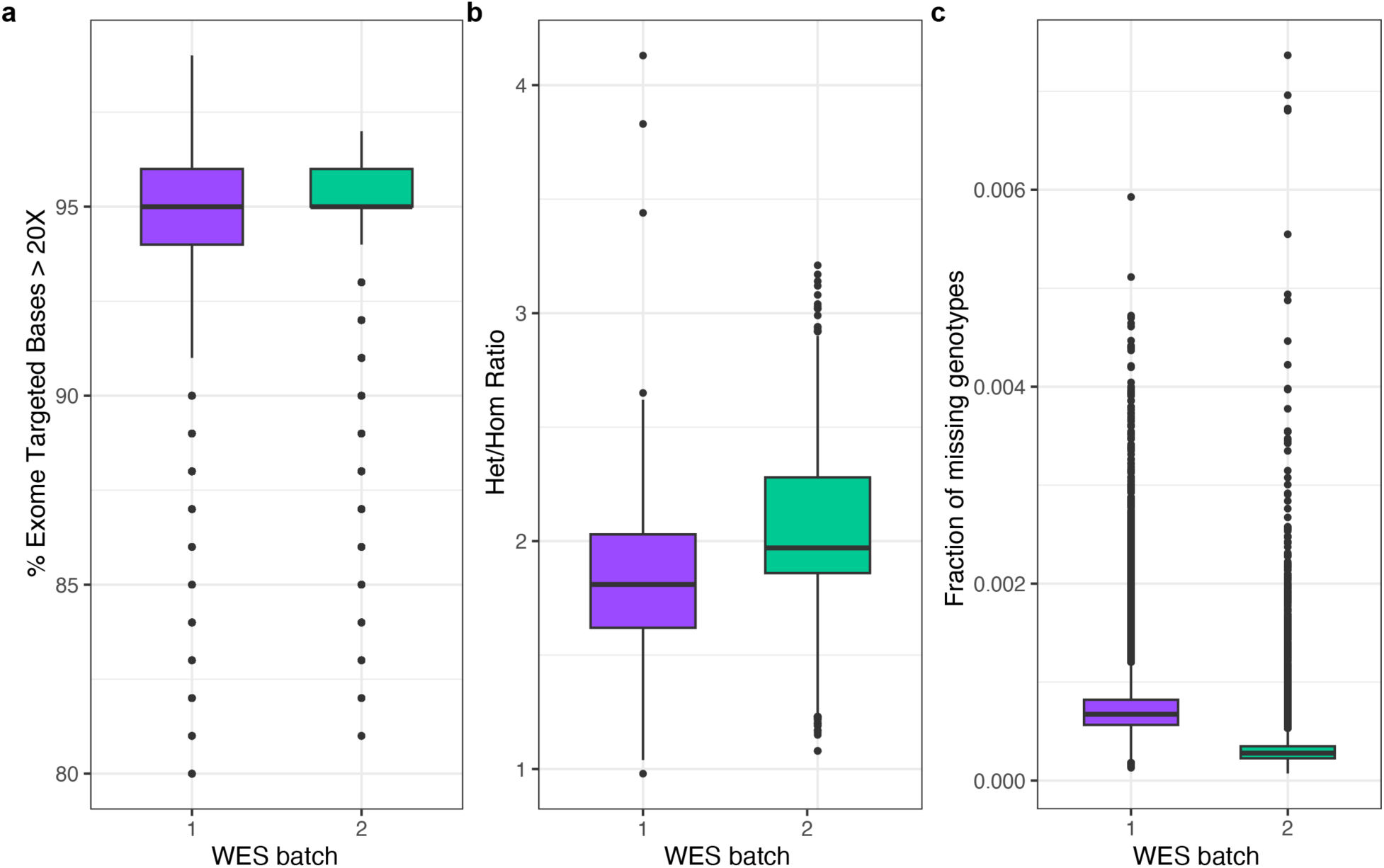
WES QC metrics. Box plots showing **a,** the percentage of exome target bases at > 20X **b,** heterozygote to homozygote ratios, and **c,** the fraction of missing genotypes for the two sequencing batches included in the study.

**Extended Data Fig. 2:**
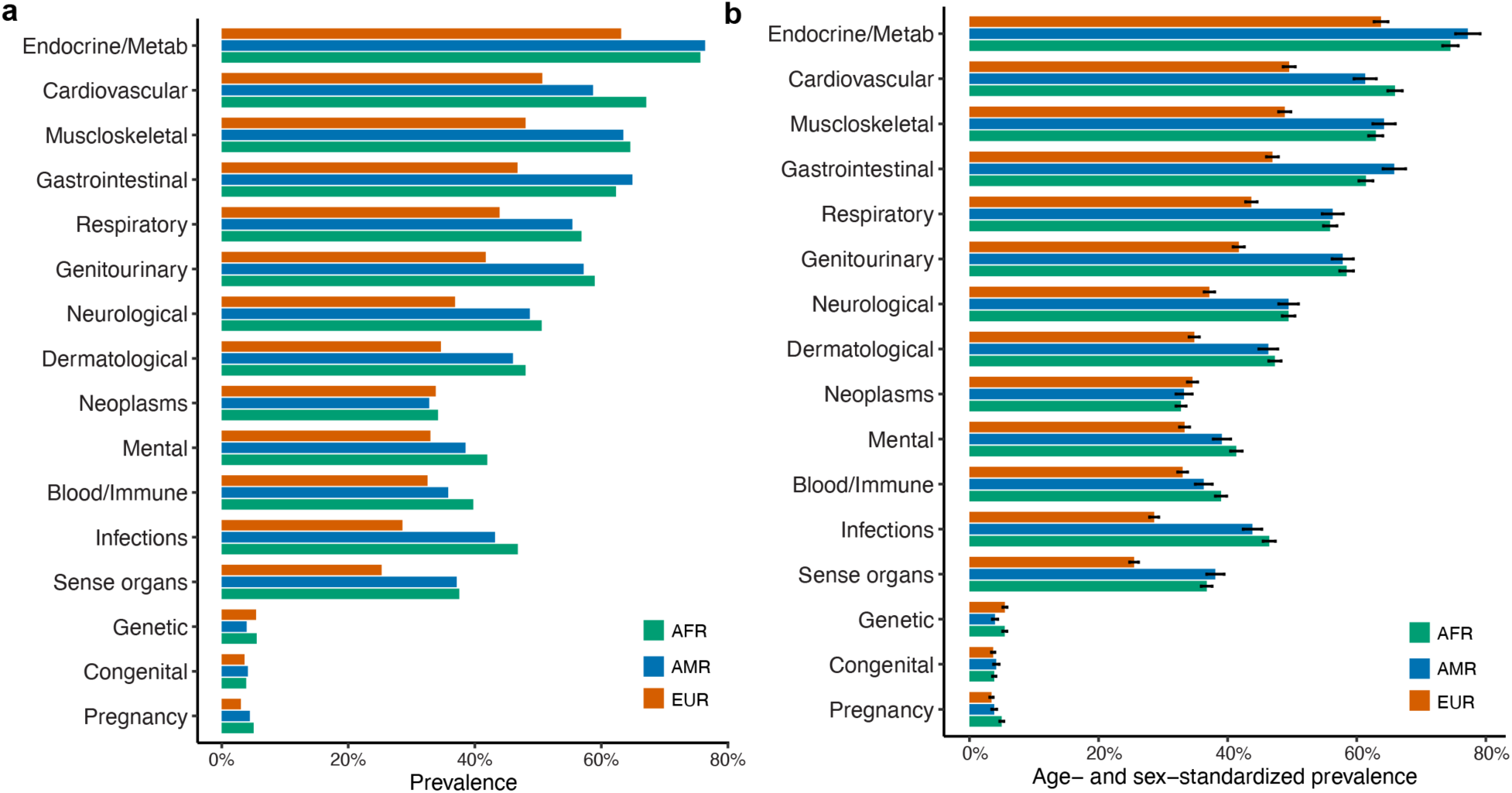
Prevalence of disease categories. Bar plots of the **a,** unstandardized and **b,** age– and sex-standardized prevalence of 16 disease categories across the three ancestry groups.

**Extended Data Fig. 3:**
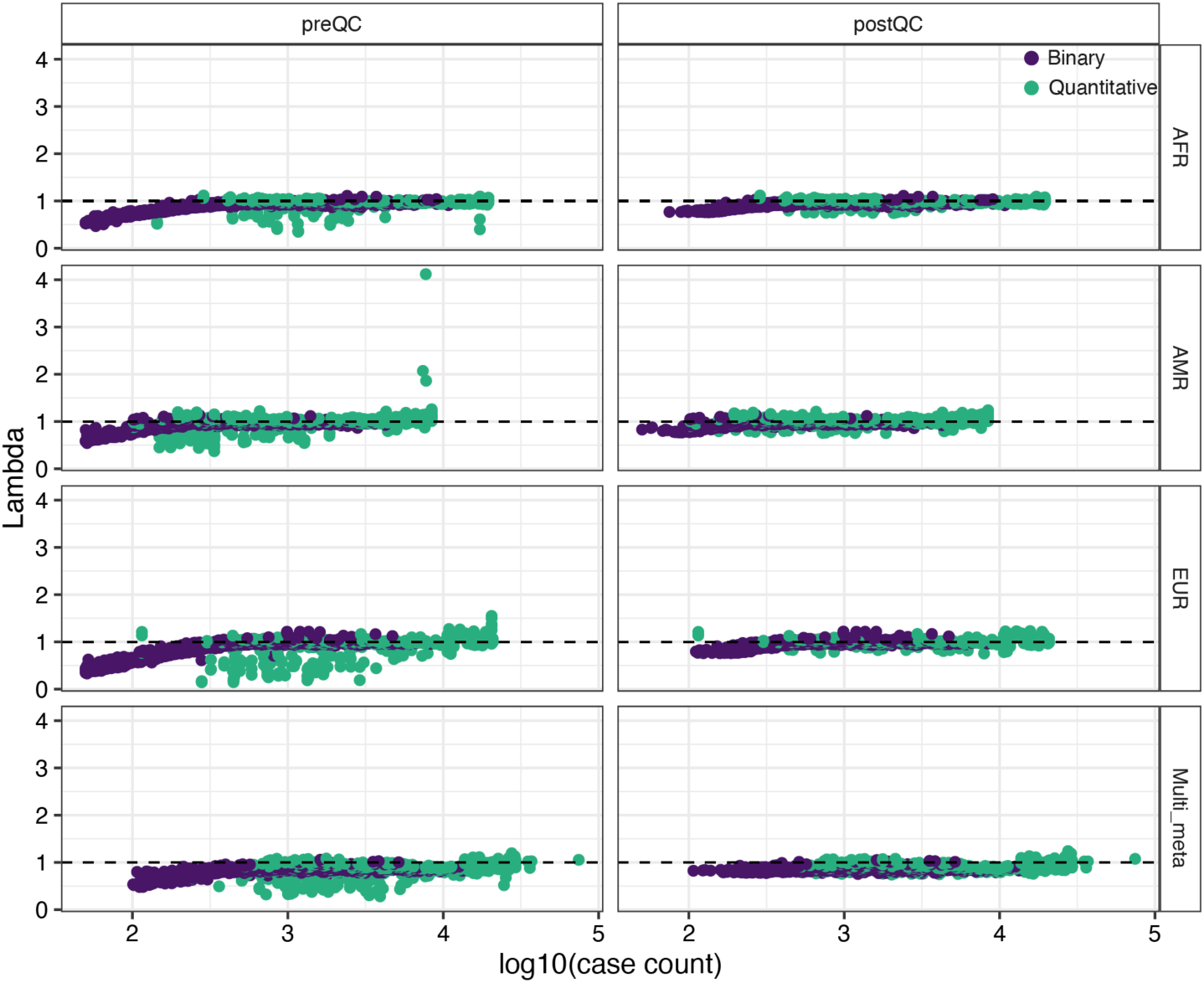
Genomic inflation factors from the gene-level PheWAS. Genomic inflation factors (lambda) were calculated for each phenotype using synonymous variants. Distributions of lambda values for binary and quantitative traits are shown before and after QC.

**Extended Data Fig. 4:**
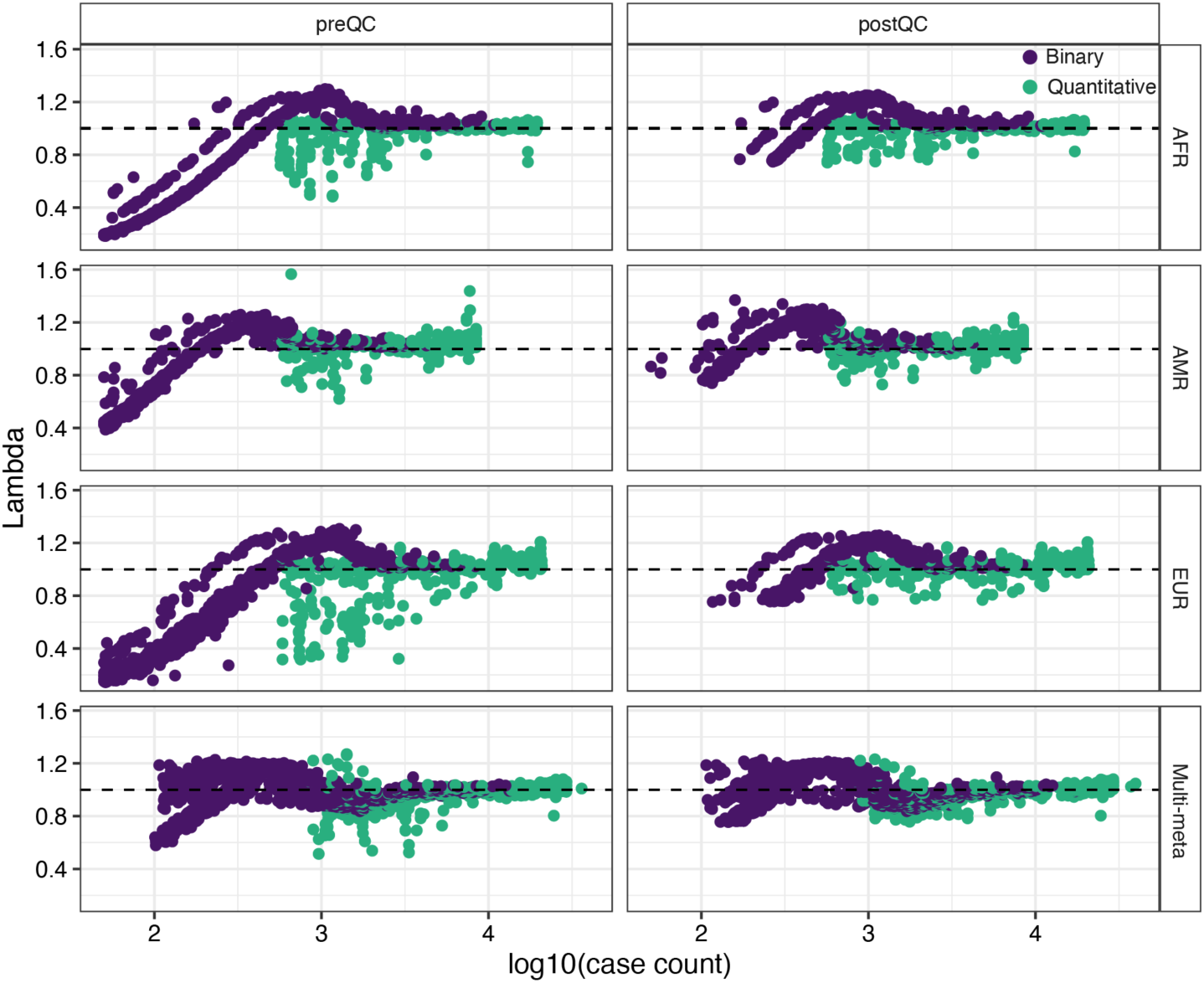
Genomic inflation factors from the variant-level PheWAS. Genomic inflation factors (lambda) were calculated for each phenotype using synonymous variants. Distributions of lambda values for binary and quantitative traits are shown before and after QC.

**Extended Data Fig. 5:**
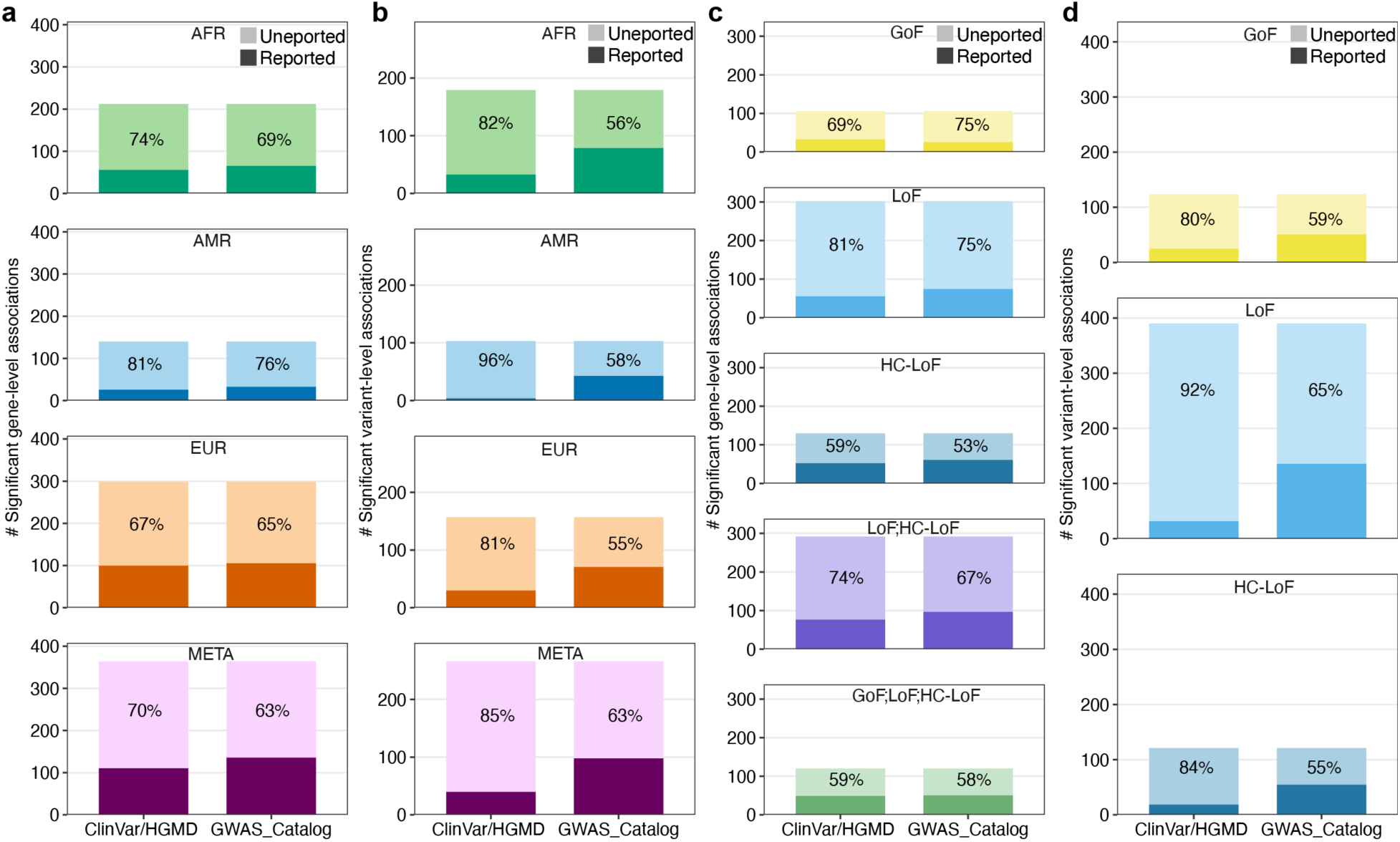
Previously reported and unreported gene-trait and variant-trait associations. Bar plots showing the proportions of previously reported and unreported associations among **a,** gene-level significant associations across ancestry-specific and multi-ancestry meta-analyses, **b,** variant-level significant associations across ancestry-specific and multi-ancestry meta-analyses, **c,** gene-level significant associations across five variant masks and **d,** variant-level significant associations across the three variant categories.

